# Chikungunya Virus VLP Vaccine: Phase 3 Trial in Adults ≥65 Years of Age

**DOI:** 10.1101/2024.10.10.24315205

**Authors:** Lauren C. Tindale, Jason S. Richardson, Debbie M. Anderson, Jason Mendy, Sufia Muhammad, Tobi Loreth, Sarah Royalty Tredo, Roshan Ramanathan, Victoria A. Jenkins, Lisa Bedell, Patrick Ajiboye, the EBSI-CV-317-005 Study Group

## Abstract

1

**Background:** Adults ≥65 years of age are at increased risk for atypical presentations of chikungunya disease as well as severe outcomes including death.

**Methods:** A phase 3, randomized, double-blind, placebo-controlled, parallel-group trial was conducted in adults ≥65 years of age where participants received a single intramuscular dose of chikungunya virus (CHIKV) virus-like particle (VLP) vaccine or placebo on Day 1. Baseline and postvaccination CHIKV serum neutralizing antibody (SNA) titers (NT_80_) were assessed at selected timepoints. Safety was assessed through Day 183.

**Results:** A total of 413 participants (206 vaccine, 207 placebo) were randomized. Coprimary endpoints were met including 1) immunologic superiority of CHIKV SNA titers compared to placebo and 2) by geometric mean titer at Day 22. CHIKV VLP vaccine induced a protective seroresponse (SNA NT_80_ ≥100; considered the presumptive seroprotective antibody response), in 82% of individuals at Day 15, in 87% of individuals at Day 22, and in 76% of individuals at Day 183. There were no notable differences in AE rates between groups and most AEs were grade 1 or 2 in severity. No vaccine-related serious adverse events or deaths occurred.

**Conclusions:** We provide robust data from adults ≥65 years of age demonstrating that CHIKV VLP vaccine has a favorable safety profile and can provide a high rate of protection within 2 weeks postvaccination and through 6 months of follow-up. (Funded by Emergent BioSolutions Inc. and Bavarian Nordic A/S [as successor in interest to Emergent BioSolutions Inc.]; ClinicalTrials.gov number, NCT05349617).

## 2 INTRODUCTION

Chikungunya virus is transmitted to humans through the bite of infected mosquitoes. In the human host, the virus incubates for 1 to 12 days before reaching high viremia levels and inducing initial symptoms of fever and flu-like syndrome.^1^ Acute febrile chikungunya cases commonly present with fever and joint pain, while other symptoms may include joint stiffness or swelling, muscle pain, lack of appetite, fatigue, headache, or rash.^2^ In nearly half of CHIKV cases, chronic symptoms such as continual or recurrent arthralgias, depression, and mood and sleep disorders persist for months or years,^3,4^ with significant impact on quality of life and daily productivity. Advanced age and severe acute symptoms are associated with worse arthritic sequelae.^5^ Additionally, advanced age has been associated with atypical (ie, neurological, cardiovascular, dermatological, ophthalmological, hepatic, renal, respiratory, hematological) and severe disease presentations when compared to younger individuals, potentially due to the increased presence of comorbidities and age-related immune system decline in older adults.^6,7^ While the overall mortality rate from chikungunya disease is low, the risk of death increases with age. A retrospective study of 127 chikungunya deaths over a 4-year period in Brazil found that while the fatality rate of chikungunya was 0.35%, the risk of death from chikungunya progressively increased starting at 40 years of age, with 14x greater risk among 40 to 49-year-olds, 28x risk among 50 to 59-year-olds, and 79x risk among individuals 60 years and older, compared to their reference age group of 20 to 29-year-olds.^8^ CHIKV vaccination is of particular importance to international travelers, both for personal disease prevention and to prevent seeding of outbreaks in nonendemic countries upon return.^9^ In a study of 1202 travelers with chikungunya from 2005-2020, individuals aged >65 years accounted for 8.5% of cases^10^ and this proportion is expected to increase with the growing population of older adults worldwide.^11,12^

CHIKV VLP vaccine (previously PXVX0317)^13^ is a formulation of 40 μg CHIKV VLP adjuvanted with aluminum hydroxide adjuvant and supplied as a pre-filled syringe for ease of use and improved dosing accuracy. Findings from studies in adults are not necessarily applicable to older adults, and it is important to assess immunogenicity and efficacy of vaccines in older age groups as it can be greatly reduced compared to younger individuals.^12^ Due to immunosenescence and the associated reduction in T cell responses that occur with aging, evaluation of the safety and effectiveness of CHIK VLP vaccine directly in older adults provides critical information on the benefits and risks in this population. Here we report safety and immunogenicity for CHIKV VLP vaccine in a robust population of older adults ≥65 years of age for up to 6 months postvaccination.

## 3 METHODS

### 3.1 Trial design, oversight, and participants

This pivotal phase 3, randomized, placebo-controlled, double-blind, parallel group trial included generally healthy participants ≥65 years of age. This was a multicenter trial conducted at 10 sites in the US. Full inclusion and exclusion criteria are provided in the Supplementary Appendix. The trial was conducted in compliance with International Council on Harmonization Good Clinical Practice guidelines and the principles in the Declaration of Helsinki. The trial protocol was approved by an independent institutional review board and by federal regulatory agencies. Written informed consent was obtained prior to any trial procedure. An independent Safety Monitoring Committee reviewed aggregated, blinded safety data after the first 50 participants completed 7 days of safety follow-up. Bavarian Nordic A/S and Emergent BioSolutions Inc. were responsible for trial design, data collection, data analysis, data interpretation, and writing of the report. Additional details regarding the design of the trial are provided in the protocol, available at NEJM.org.

### 3.2 Randomisation and masking

Participants were stratified into two age groups (65 to 74 and ≥75 years of age) and randomized in a 1:1 ratio to receive either CHIKV VLP vaccine or placebo; 400 participants were planned for enrollment. Participants attended a screening visit, then a Day 1 visit which included randomization, baseline blood collection, and administration of a single dose of CHIKV VLP vaccine or placebo by intramuscular injection in the deltoid muscle (Figure S1). Day 15, 22, and 183 visits included review of adverse events (AEs) and concomitant medications and blood collection for immunogenicity analysis. Day 29 and 92 phone visits included review of AEs.

### 3.3 Immunogenicity

A luciferase-based CHIKV human SNA assay was used to measure 80% serum neutralizing antibody (SNA) titer (NT_80_) against CHIKV in human serum samples (Supplementary Appendix). Seroresponse rate (SRR) was defined as the percentage of participants who achieved SNA NT_80_ ≥100 (presumptive seroprotection rate agreed with the FDA and EMA).

The coprimary endpoints were: 1) difference in CHIKV SNA SRR (vaccine minus placebo) at Day 22, and 2) CHIKV SNA geometric mean titer (GMT) at Day 22 for vaccine and placebo.

The key secondary endpoints were the difference in CHIKV SNA SRR at Day 15 and Day 183, in that order. Other secondary endpoints included 1) CHIKV SNA GMTs by trial arm at Day 15 and Day 183, 2) geometric mean fold increase (GMFI) from Day 1 to subsequent collection time points, and 3) number and percentage of participants with SNA NT_80_ ≥15, and 4-fold rise over baseline.

### 3.4 Safety

Participants were monitored for signs of an acute adverse reaction(s) to vaccination for 30 minutes. Solicited AEs were collected from injection administration on Day 1 through Day 8 using a diary. Solicited AEs included local events of pain, redness, and swelling at the injection site and systemic events of oral temperature ≥38.0°C (≥100.4°F), chills, fatigue, headache, myalgia, arthralgia, and nausea. Unsolicited AEs were collected from Day 1 through Day 29. Serious AEs (SAEs), AEs of special interest (AESI) (defined as new onset or worsening arthralgia that was medically attended), and medically attended AEs (MAAEs) were collected from Day 1 postvaccination through Day 183. The investigator graded all AEs for severity as grade 1 (mild), grade 2 (moderate), grade 3 (severe), or grade 4 (potentially life-threatening). The investigator assessed AE causality. If the relationship between the AE and the trial drug was determined to be “possible” or “probable,” the event was considered “related.”

### 3.5 Statistical analysis

The sample size was chosen based on expected SRR to be approximately 90% vs <5% for the placebo participants at Day 22. With an assumed 10% rate of nonevaluable participants, the power to show superiority over placebo with 180 CHIKV VLP vaccine and 180 placebo evaluable participants was >99.9% for the combined age groups. The difference in SRR rate between vaccine and placebo groups that was considered clinically relevant was ≥70%. With 180 baseline seronegative vaccine treated participants and a target SRR of 90% vs a rate of 5% for placebo, the width of a two-sided 95% CI would be ±5.4%. If the target vaccine seroresponse was 77%, the width would be ±6.9%. Therefore, the difference in SRR had to be above 77% for the lower bound of the 95% CI for the difference to be ≥70%.

The exposed population included all participants who received a CHIKV VLP vaccine or placebo injection. The safety population included all participants from the exposed population who provided safety assessment data. The modified intent-to-treat (mITT) population included all participants who were vaccinated and had at least one post-injection CHIKV SNA result. The immunogenicity evaluable population (IEP) included mITT population participants who had no measurable CHIKV SNA at Day 1 (below LLOQ), had evaluable SNA at Day 22, and had no reason for exclusion as defined prior to unblinding (primary population for immunogenicity analysis). Additional details are provided in the statistical analysis plan, available at NEJM.org.

The primary objectives of the trial were based on SNA NT_80_ titers calculated to determine the CHIKV neutralizing antibody response. The difference between treatment groups in SRR (proportion of participants with SNA NT_80_ ≥100) was tested using a chi-square test with a 2-sided alpha=0.05 in the IEP across both age groups combined and the 2-sided 95% CI for the difference was computed using the Newcombe hybrid score method. In the computation of the GMTs, values below the assay lower limit of quantitation (LLOQ) of <15 were assigned the value LLOQ/2=7.5. The GMTs were compared between CHIKV VLP vaccine and placebo treatment groups and were analyzed via a linear model based on a 2-sided alpha=0.05. The primary model was an ANOVA, with logarithmically transformed CHIKV SNA titers (log_10_) as the dependent variable and treatment group and trial site as the fixed effects in the model. The adjusted least square means and their 95% CIs calculated based on the ANOVA were back transformed and reported as the group GMT values. The proportion of participants with seropositivity (SNA NT_80_ ≥15; above the LLOQ) and those with a 4-fold increase in titer over baseline were also summarized. Multiplicity was addressed by requiring both coprimary endpoints to be met as a success criterion and via hierarchical testing for the key secondary endpoints.

Safety data, frequency and percentage of participants with either solicited or unsolicited AEs, were summarized descriptively including duration, severity, and causality of the events observed.

## 4 RESULTS

### 4.1 Participants

Between May 12, 2022, and December 2, 2022, a total of 594 volunteers were screened for this trial, of which 413 were eligible, randomized, and administered investigational product. The IEP included 372 participants, of which 189 received CHIKV VLP vaccine and 183 received placebo (**Figure 1**).

**Figure 1:**
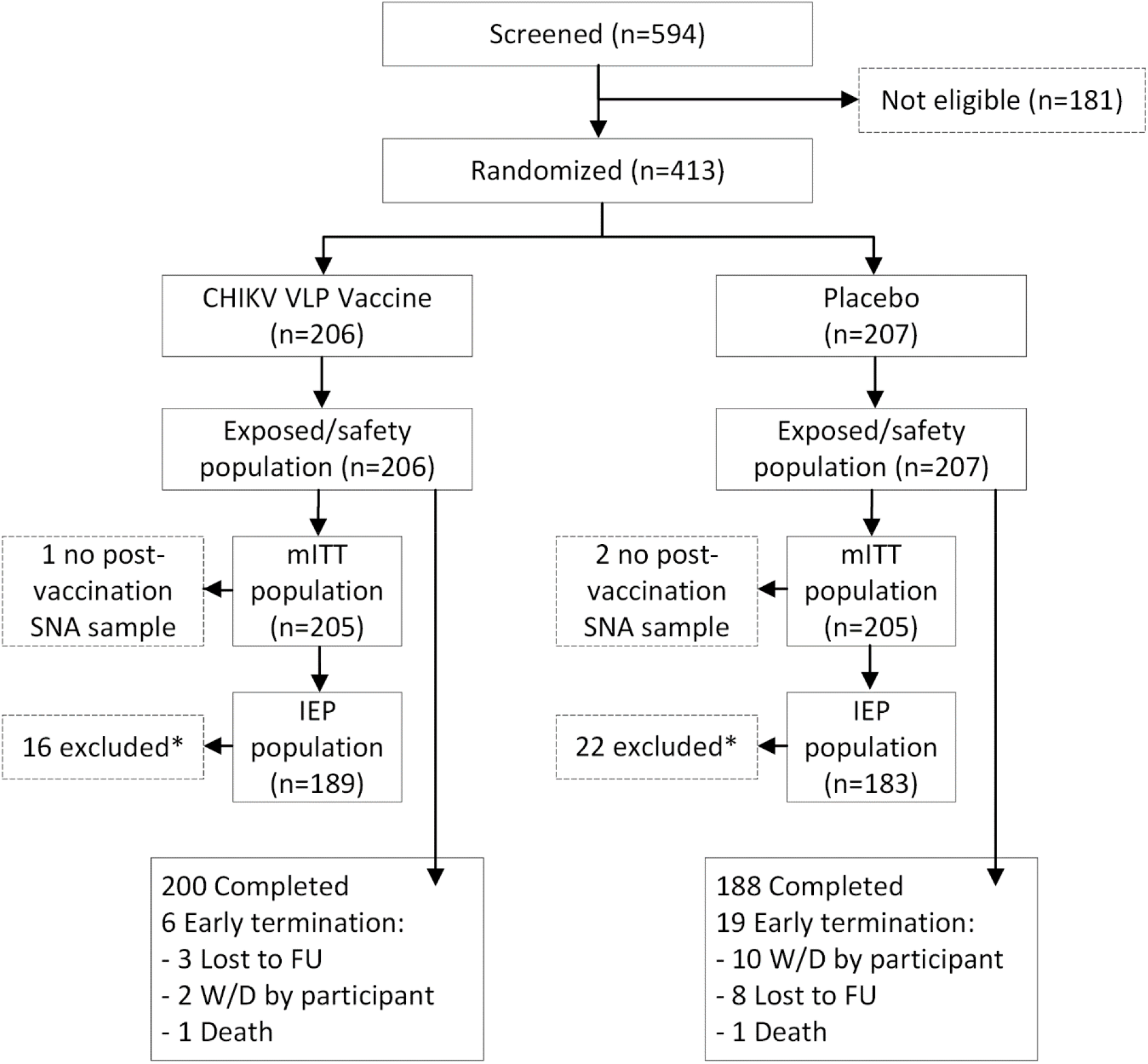
Trial profile. *Exclusions from IEP were due to: no SNA result in Day 22 window, measurable Day 1 SNA (baseline positive), prohibited vaccine/medication use, eligibility criteria not met, missing Day 1 SNA sample, and/or serum processing error. FU = follow up; IEP = immunogenicity evaluable population; LLOQ = lower limit of quantitation; mITT = modified intent-to-treat; SNA = serum neutralizing antibody; W/D = withdrawn.

There was no appreciable difference between the CHIKV VLP vaccine and placebo groups in demographic characteristics (**Table 1**). In the randomized population, 39.3% of participants were male. There were 318 participants aged 65 to 74 (159 vaccine, 159 placebo) and 95 participants aged ≥75 years (47 vaccine, 48 placebo). White participants accounted for 83.3% of randomized population and 11.9% were Black or African American.

**Table 1:**
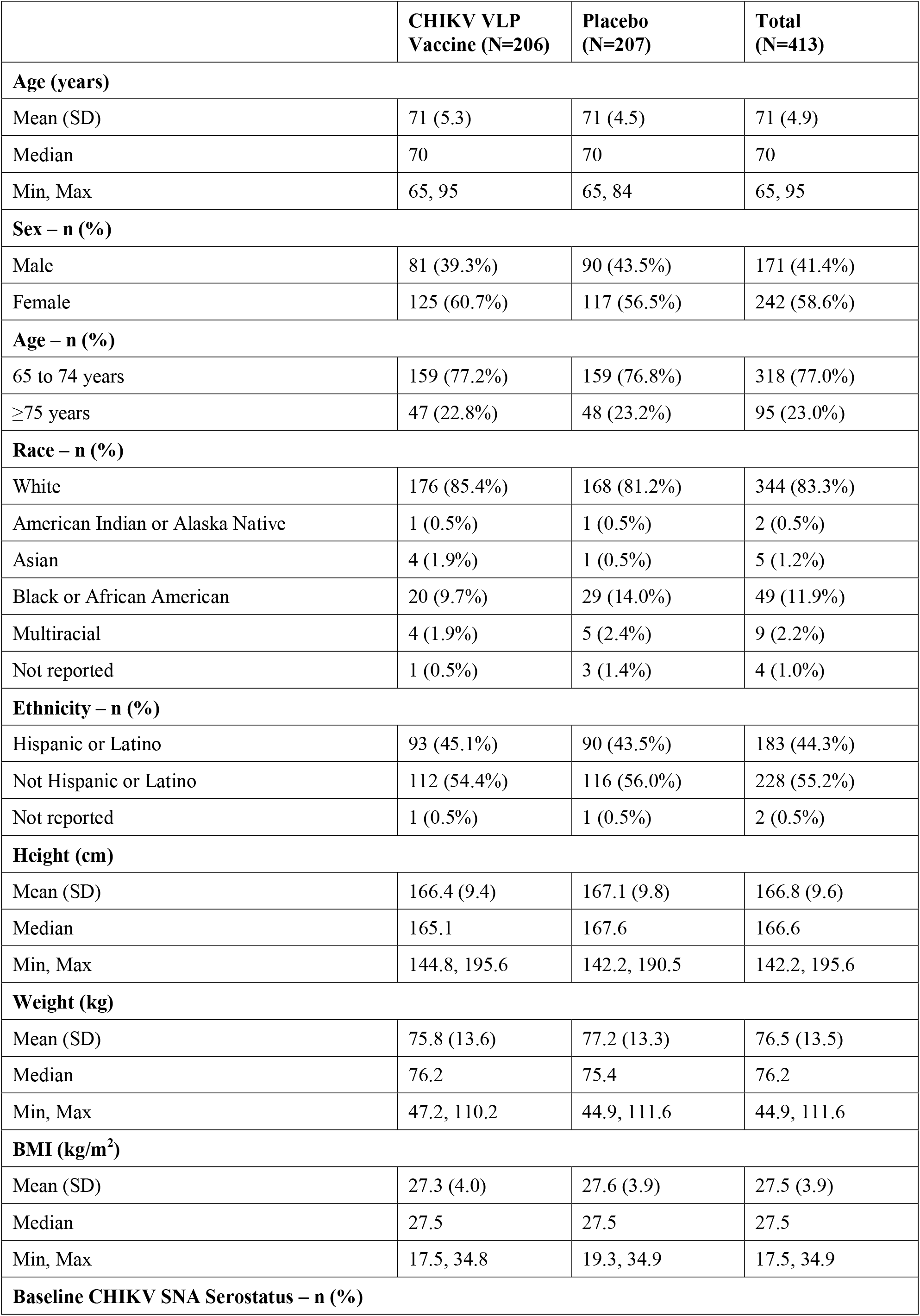
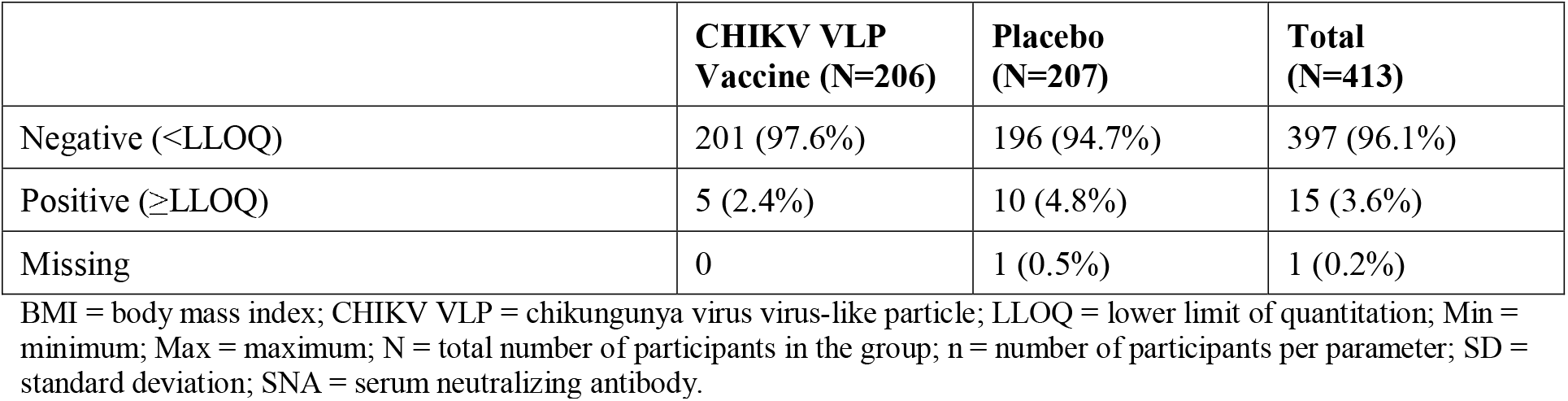
Baseline Characteristics of the Randomized Population.

|  | **CHIKV VLP**<br>**Vaccine (N=206)** | **Placebo**<br>**(N=207)** | **Total**<br>**(N=413)** |
| --- | --- | --- | --- |
| **Age (years)** |  |  |  |
| Mean (SD) | 71 (5.3) | 71 (4.5) | 71 (4.9) |
| Median | 70 | 70 | 70 |
| Min, Max | 65, 95 | 65, 84 | 65, 95 |
| **Sex – n (%)** |  |  |  |
| Male | 81 (39.3%) | 90 (43.5%) | 171 (41.4%) |
| Female | 125 (60.7%) | 117 (56.5%) | 242 (58.6%) |
| **Age – n (%)** |  |  |  |
| 65 to 74 years | 159 (77.2%) | 159 (76.8%) | 318 (77.0%) |
| ≥75 years | 47 (22.8%) | 48 (23.2%) | 95 (23.0%) |
| **Race – n (%)** |  |  |  |
| White | 176 (85.4%) | 168 (81.2%) | 344 (83.3%) |
| American Indian or Alaska Native | 1 (0.5%) | 1 (0.5%) | 2 (0.5%) |
| Asian | 4 (1.9%) | 1 (0.5%) | 5 (1.2%) |
| Black or African American | 20 (9.7%) | 29 (14.0%) | 49 (11.9%) |
| Multiracial | 4 (1.9%) | 5 (2.4%) | 9 (2.2%) |
| Not reported | 1 (0.5%) | 3 (1.4%) | 4 (1.0%) |
| **Ethnicity – n (%)** |  |  |  |
| Hispanic or Latino | 93 (45.1%) | 90 (43.5%) | 183 (44.3%) |
| Not Hispanic or Latino | 112 (54.4%) | 116 (56.0%) | 228 (55.2%) |
| Not reported | 1 (0.5%) | 1 (0.5%) | 2 (0.5%) |
| **Height (cm)** |  |  |  |
| Mean (SD) | 166.4 (9.4) | 167.1 (9.8) | 166.8 (9.6) |
| Median | 165.1 | 167.6 | 166.6 |
| Min, Max | 144.8, 195.6 | 142.2, 190.5 | 142.2, 195.6 |
| **Weight (kg)** |  |  |  |
| Mean (SD) | 75.8 (13.6) | 77.2 (13.3) | 76.5 (13.5) |
| Median | 76.2 | 75.4 | 76.2 |
| Min, Max | 47.2, 110.2 | 44.9, 111.6 | 44.9, 111.6 |
| **BMI (kg/m<sup>2</sup>)** |  |  |  |
| Mean (SD) | 27.3 (4.0) | 27.6 (3.9) | 27.5 (3.9) |
| Median | 27.5 | 27.5 | 27.5 |
| Min, Max | 17.5, 34.8 | 19.3, 34.9 | 17.5, 34.9 |
| **Baseline CHIKV SNA Serostatus – n (%)** |  |  |  |

**Table 1: Baseline Characteristics of the Randomized Population.**
|  | **CHIKV VLP Vaccine (N=206)** | **Placebo (N=207)** | **Total (N=413)** |
| --- | --- | --- | --- |
| Negative (<LLOQ) | 201 (97.6%) | 196 (94.7%) | 397 (96.1%) |
| Positive (≥LLOQ) | 5 (2.4%) | 10 (4.8%) | 15 (3.6%) |
| Missing | 0 | 1 (0.5%) | 1 (0.2%) |

BMI = body mass index; CHIKV VLP = chikungunya virus virus-like particle; LLOQ = lower limit of quantitation; Min = minimum; Max = maximum; N = total number of participants in the group; n = number of participants per parameter; SD = standard deviation; SNA = serum neutralizing antibody.

### 4.2 Immunogenicity

At Day 22, 87.3% (165/189) of participants in the CHIKV VLP vaccine group had a seroresponse (SNA NT_80_ ≥100), compared to 1.1% (2/183) of participants in the placebo group for the IEP (**Table 2**). The difference in SRR was 86.2% (95% CI: 80.0%, 90.3%) which was statistically significant (P<0.0001, chi-square test) and clinically relevant (lower bound of the 95% CI ≥70%). CHIKV SNA GMT at Day 22 was 724 for the vaccine group and 8 for the placebo group, for a GMT ratio of 90 (95% CI: 69, 117; P<0.0001, ANOVA) for the IEP (Table S1 in the Supplementary Appendix). All coprimary endpoints were satisfied.

**Table 2:**
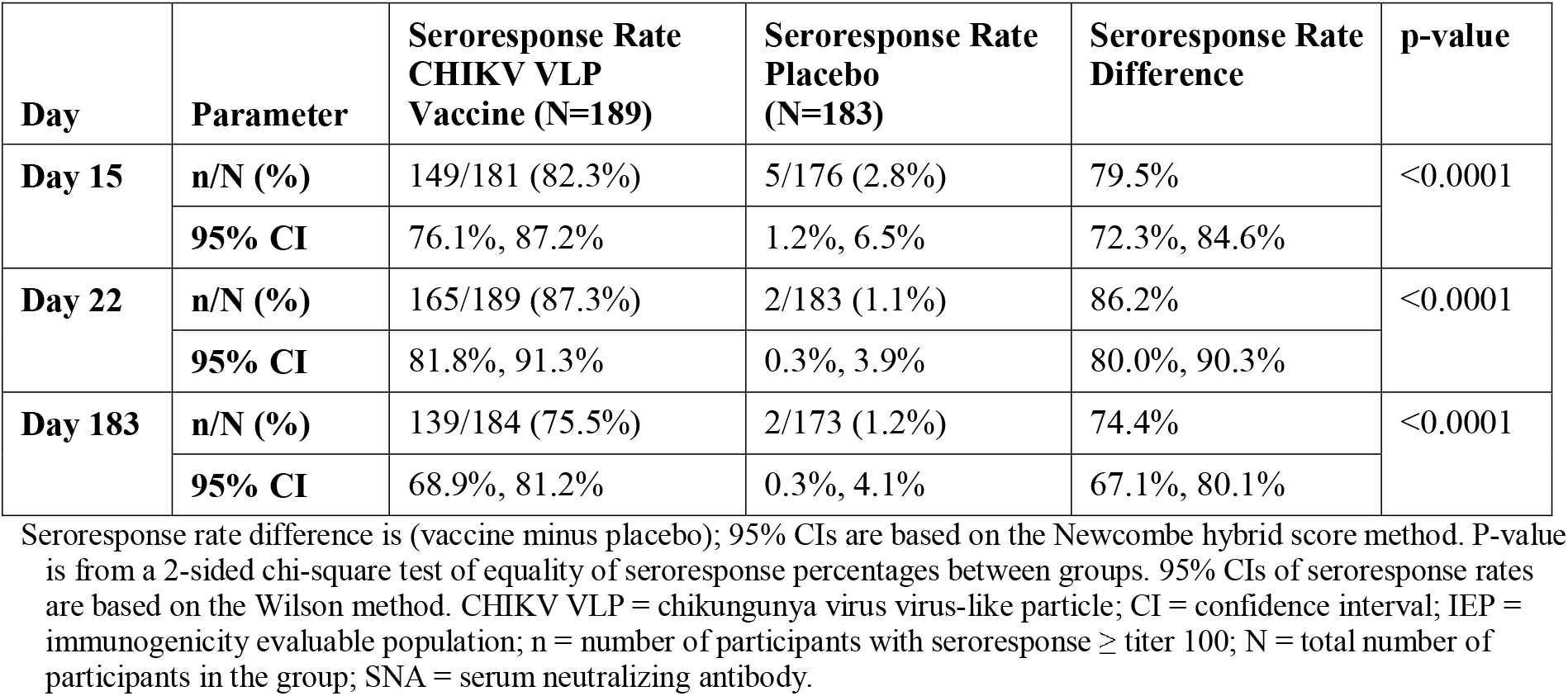
CHIKV Serum Neutralizing Antibody Seroresponse Rate and Seroresponse Rate Difference by Trial Visit (Immunogenicity Evaluable Population)

At Day 15, 82.3% of participants in the CHIKV VLP vaccine group had a seroresponse, compared to 2.8% participants in the placebo group. The SRR difference at Day 15 was 79.5% [95% CI: 72.3%, 84.6%], which was statistically significant (P<0.0001, chi-square test) and clinically relevant (lower bound of the 95% CI ≥70%). At Day 183, 75.5% of participants in the vaccine group had a seroresponse, compared to 1.2% in the placebo group (P<0.0001, chi-square test).

Both SNA NT_80_ GMT and GMFI were significantly higher in the CHIKV VLP vaccine group than the placebo group at all trial days after vaccination. In the vaccine group, the SNA GMT increased from below the LLOQ at Day 1 to a titer of 378 at Day 15, followed by a rise to a peak titer of 724 at Day 22, and then decreased at Day 183 to 233. This corresponded to a GMFI of 49 at Day 15, 94 at Day 22, and 29 at Day 183. Nearly all participants had a measurable SNA response to CHIKV VLP vaccine. Seroconversion (SNA NT_80_ ≥15) was seen in 94.5% of vaccine recipients at Day 15, 95.2% at Day 22, and 92.9% at Day 183. Placebo group participants who had a seroresponse (n=9) were investigated and identified as potential sample handling errors, as each of these participants had SNA NT_80_ ≥100 result at a single timepoint only, with SNA NT_80_ <15 at all other sample collection times.

By age group, the SRR for Day 15 showed that the younger age group elicited a more rapid response (**Figure 2A**, Table S2 in the Supplementary Appendix). In the 65 to 74 age group, the SRR for the vaccine group was 84.0% at Day 15, 87.9% at Day 22, and 76.2% at Day 183, compared to 75.7% at Day 15, 85.0% at Day 22 and 73.0% at Day 183 in the ≥75 age group. The GMT at Day 15 was higher in the 65 to 74 age group (407) compared to the ≥75 age group (299), at Day 22 the GMTs were comparable (65 to 74 age group: 726; ≥75 age group: 716), and at Day 183 the GMTs were slightly higher in the older age group (65 to 74 age group: 222; ≥75 age group: 281) (**Figure 2B**).

**Figure 2:**
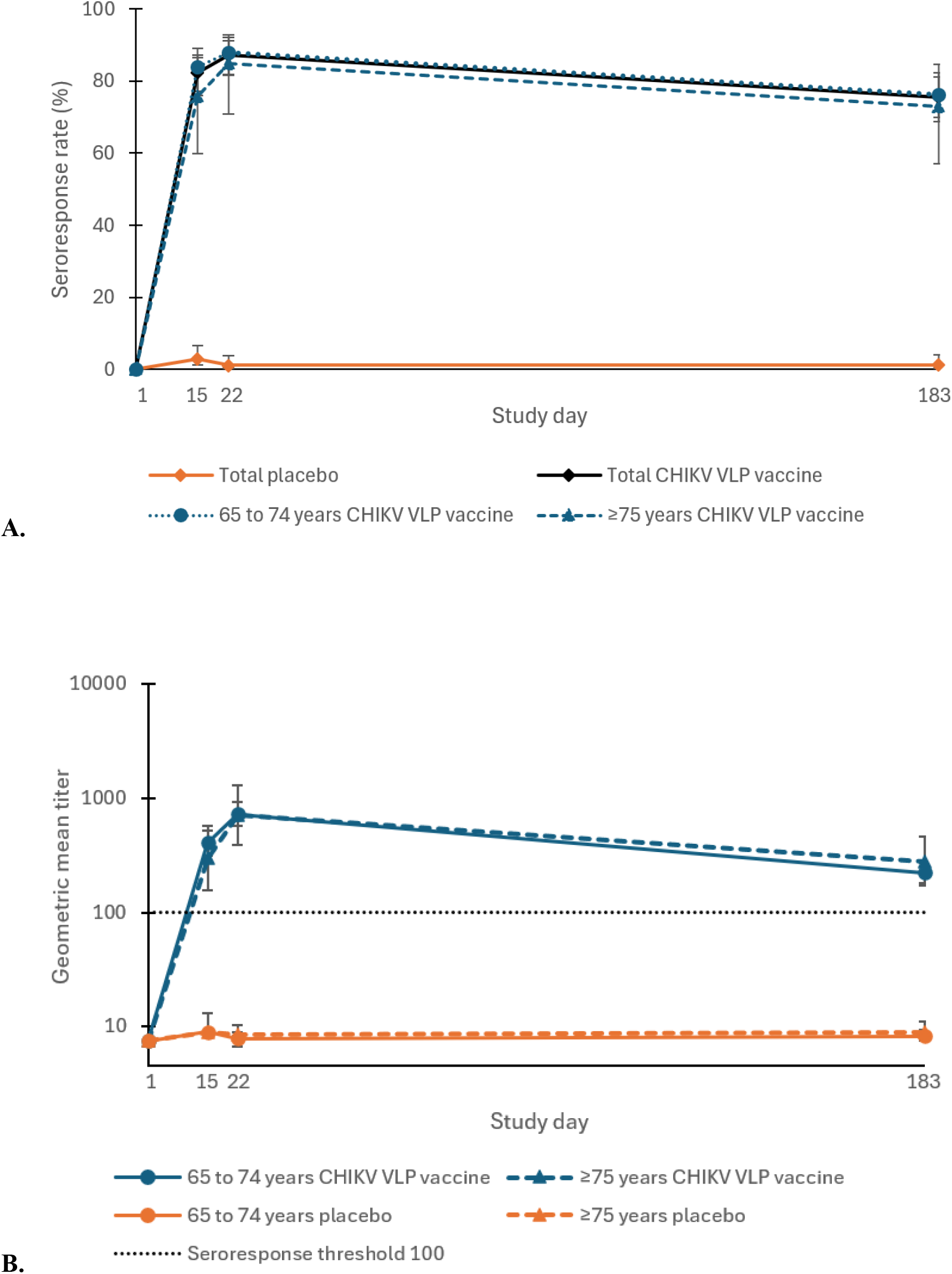
A. Seroresponse Rate and B. Geometric Mean Titers for CHIKV Serum Neutralizing Antibodies by Visit and by Age Group (Immunogenicity Evaluable Population). Y-axis is represented on a log_10_ scale. Vertical bars denote the 95% confidence interval. Values below lower limit of quantitation (LLOQ=15) were assigned the value LLOQ/2=7.5.

### 4.3 Safety

A total of 22.8% of participants who received the CHIKV VLP vaccine experienced an AE compared to 25.1% in the placebo group (**Table 3**). The participants with AEs that were ≥grade 3 was 1.9% in the CHIKV VLP vaccine group and 1.4% in the placebo group. A total of 25 (12.2%) vaccine and 28 (14.0%) placebo group participants reported a solicited AE; a single participant in the vaccine group experienced 2 grade 3 solicited AEs (headache and fatigue). The most reported systemic solicited AEs were myalgia (vaccine:6.3%; placebo: 6.5%) and fatigue (vaccine: 6.3%; placebo 6.0%), each occurring in more than 5% of participants. The most reported local solicited AE was injection site pain, reported by 11 participants (5.4%) in the vaccine group and 3 participants (1.5%) in the placebo group. Unsolicited AEs were experienced by 26 (12.6%) participants in the CHIKV VLP vaccine group and 34 (16.4%) in the placebo group, with 4 (1.9%) and 6 (2.9%) participants having treatment-related unsolicited AEs, respectively. There was one treatment-related unsolicited AE that was ≥grade 3 (vaccine group, grade 3 fatigue).

**Table 3:** Summary of Adverse Events (Safety Population)

|  | **CHIKV VLP Vaccine**<br>**(N=206) n (%)** | **Placebo**<br>**(N=207) n (%)** |
| --- | --- | --- |
| Any adverse event (solicited and unsolicited) | 47 (22.8) | 52 (25.1) |
| ≥ Grade 3 | 4 (1.9) | 3 (1.4) |
| Any unsolicited adverse event | 26 (12.6) | 34 (16.4) |
| ≥ Grade 3 | 4 (1.9) | 3 (1.4) |
| Any related unsolicited adverse event | 4 (1.9) | 6 (2.9) |
| ≥ Grade 3 | 1 (0.5) | 0 |
| Any serious adverse event | 4 (1.9) | 3 (1.4) |
| Any related serious adverse event | 0 | 0 |
| Any adverse event of special interest | 0 | 1 (0.5) |
| Any related adverse event of special interest | 0 | 0 |
| Any fatal adverse event | 1 (0.5) | 1 (0.5) |
| Any local solicited adverse event | 11 (5.4) | 4 (2.0) |
| ≥ Grade 3 | 0 | 0 |
| Injection site pain | 11 (5.4) | 3 (1.5) |
| Injection site redness | 0 | 1 (0.5) |
| Injection site swelling | 0 | 0 |
| Any systemic solicited adverse event | 22 (10.7) | 27 (13.5) |
| ≥ Grade 3 | 1 (0.5) | 0 |
| Myalgia | 13 (6.3) | 13 (6.5) |
| Fatigue | 13 (6.3) | 12 (6.0) |
| Headache | 9 (4.4) | 15 (7.5) |
| Arthralgia | 6 (2.9) | 8 (4.0) |
| Chills | 6 (2.9) | 6 (3.0) |
| Nausea | 6 (2.9) | 3 (1.5) |
| Fever | 0 | 2 (1.0) |

Relationship is investigator's assessment. Percentages are based on the number of safety population participants in each treatment group. Participants are counted at most once in each category. Percentages for solicited events are based on the number of participants with at least one diary observation (excluding 'Not done/unknown') for a given symptom or a given day. CHIKV VLP = chikungunya virus virus-like particle; N = total number of participants in the group; n = number of participants with events.

There were 4 (1.9%) participants who had an SAE in the vaccine group and 3 (1.4%) in the placebo group, none of which were treatment related. A single participant experienced an AESI (0.5%), defined as new onset or worsening arthralgia that was medically attended, in the placebo group (grade 2 joint dislocation). Treatment-related MAAEs were experienced by 2 participants in the vaccine group (grade 3 fatigue and grade 1 hypertension). There were 2 fatal SAEs, 1 in the vaccine group and 1 in the placebo group, neither of which were treatment-related (respiratory failure and lung cancer).

## 5 DISCUSSION

Immunosenescence, the progressive age-related decline of the immune system, is associated with increased susceptibility, severity, and mortality from infectious diseases as well as a diminished response to vaccination in older adults.^14^ In this pivotal phase 3 trial, we provide results from in a robust population of ≥65-year-old individuals. CHIKV VLP vaccine was immunogenic in this critical age group as demonstrated by 87.3% of participants reaching seroprotective levels of antibodies and vaccine superiority to placebo in GMTs at Day 22 (21 days postvaccination). CHIKV VLP vaccine also produced early seroprotective antibody levels in 82.3% of participants at Day 15 (14 days postvaccination) and persisted through Day 183 (6 months postvaccination) of follow-up in 75.5% of participants. A subgroup analysis of immune responses to CHIKV VLP vaccine by age subgroups: (65 to 74 years and ≥75 years of age) was performed. While there was a slightly higher early immune response in the 65 to 74 age group at Day 15 (SRR = 84.0%) compared to the ≥75 age group (SRR = 75.7%), the seroresponse rates at Day 22 (SRR = 87.9% and 85.0%, respectively) and at Day 183 (SRR = 76.2% and 73.0%, respectively) were comparable.

The CHIKV VLP vaccine had a favorable safety profile, with most AEs being self-limited and grade 1 to 2 in severity. There were no notable differences in solicited or unsolicited AE rates between treatment groups. The most frequently reported systemic solicited AEs were myalgia and fatigue, each occurring in more than 5% of participants in both the vaccine and placebo groups. The most common local solicited AE was injection site pain. On average, duration of symptoms for treatment-related solicited AEs in the CHIKV VLP vaccine group was 2.1 days with Day 2 being the most common day of symptom onset. Serious AEs were infrequent (<2%) with none considered related to investigational product. The incidence of AESI and MAAEs did not differ between the CHIKV VLP vaccine group and the placebo group.

As the older adult population continues to grow worldwide, it is important that travel medicine targets these travelers for vaccination programs as immunosenescence and comorbidities increase their risk of morbidity and mortality from infectious diseases.^11,12^ The US Advisory Committee on Immunization Practices (ACIP) recognizes the increased need for chikungunya disease protection in older adults and while they recommend chikungunya vaccination for persons aged ≥18 years traveling to a country or territory where there is a CHIKV outbreak, they also provide additional recommendations for persons aged >65 years traveling to a country or territory without an outbreak but with evidence of CHIKV transmission among humans within the last 5 years, particularly those with underlying medical conditions. Indeed, risk factors for hospitalization include very young age and higher age as well as certain preexisting comorbidities including cardiovascular or respiratory conditions, hypertension, and diabetes.^15,16^ There is a growing global population of older adults and vaccination is a critical approach to protect against chikungunya disease.^12^

Virus-like particle vaccines have several advantages over conventional vaccines (live-attenuated and inactivated vaccines) as VLP structure closely mimics the structure of the native virus, VLP vaccines tend to be immunogenic at lower doses, and VLPs do not replicate, making them generally safe, particularly in populations for whom live-attenuated vaccines have been contraindicated (eg, pregnant women and immunocompromised).^17,18^ CHIKV VLP vaccine provides a safe and immunogenic vaccine option.

A limitation of this trial is that the determination of CHIKV VLP vaccine effectiveness was based on an immunological surrogate-marker. The immunogenicity threshold used in this trial was defined based a nonhuman primate passive antibody transfer study where human sera from CHIKV VLP vaccine vaccinated individuals was used to prevent viremia following wild-type CHIKV challenge in cynomolgus macaques. In nonhuman primates, an SNA titer of 23.6 was estimated to be associated with an 80% probability of protection from CHIKV. Based on those results, a conservative threshold of SNA NT_80_=100 was selected as the protective threshold in humans (FDA/EMA agreed threshold). Efficacy against CHIKV disease will be investigated in a future clinical endpoint trial to be conducted in CHIKV endemic regions. Another limitation is that the trial population included healthy individuals; further trials are needed to assess specific disease populations. Additional trials are currently underway to assess CHIKV VLP vaccine long-term safety and a booster dose (NCT06007183).

CHIKV VLP vaccine is the only VLP-based vaccine option with a proposed indication for active immunization to prevent disease caused by CHIKV infection in individuals aged 12 to 64 years (NCT05072080) and ≥65 years. Findings from this pivotal phase 3 trial provide robust data in older adults ≥65 years of age that demonstrate that the CHIKV VLP platform delivers a nonreplicating, safe, and immunogenic chikungunya vaccine option for a wide range of travelers. CHIKV VLP vaccine induces a rapid and robust immune response in most individuals by 14 days postvaccination, with protection lasting through 6-months.

## Data Availability

De-identified data produced in the present study are available upon reasonable request to the authors.

## 6 ACKNOWLEDGEMENTS

We thank all participants, investigators, and trial site personnel who took part in this clinical trial. We also thank all employees of Bavarian Nordic A/S and Emergent BioSolutions Inc. past and present who were involved, as well as members of the Safety Monitoring Committee.

## Notes

### Competing Interest Statement

All authors are current or previous employees/contractors of Bavarian Nordic and/or Emergent BioSolutions.

### Clinical Trial

NCT05349617

### Funding Statement

This study was funded by Emergent BioSolutions Inc. and Bavarian Nordic A/S [as successor in interest to Emergent BioSolutions Inc.]

### Author Declarations

WCG (Western-Copernicus Group) Institutional Review Board gave ethical approval for this work.

### Summary of Updates

Table 2 was revised with the correct "N" in the column header.

