## Supplementary Appendix for "Chikungunya Virus VLP Vaccine: Phase 3 Trial in Adults ≥65 Years of Age"

Dr. John E. Ervin, AMR Kansas City, Kansas City, MO

Dr. Cayce Tangeman, Coastal Carolina Research Center, North Charleston, SC

Dr. Matthew Davis, Rochester Clinical Research, Inc., Rochester, NY

Dr. Vicki Miller, DM Clinical Research, Tomball Tomball, TX

Dr. Ramon Reyes, BHFC Research, San Antonio, TX

Dr. Jorge Caso, Suncoast Research Associates, LLC, Miami, FL

Dr. Jennifer Boston, Spaulding Clinical, West Bend, WI

Dr. Robert Perry, Panax Clinical Research, Miami Lakes, FL

Dr. Lawrence Galitz, Global Clinical Research Professionals, Saint Petersburg, FL

Dr. Muhammad Ifran, DM Clinical Research CyFair, Houston, TX

Safety Monitoring Committee

Chairs

Mark S. Riddle, DrPH, MD, MPH&TM (until 31‑Oct‑2022)

Pfizer Vaccines Research and Development and University of Nevada, Reno School of Medicine

David R. Snydman, MD, FACP, FIDSA, FAST (Jan-2023 onwards)

Tufts Medical Center and Tufts University School of Medicine

Members

Michelle Petri, MD MPH

Johns Hopkins University School of Medicine and Johns Hopkins Hospital

Ying Lu, PhD

Stanford University School of Medicine

Supplementary methods

Inclusion and exclusion criteria

Inclusion Criteria

1. Able and willing to provide informed consent voluntarily signed by participant. Must verbalize understanding of the general procedures of, and reason for the study.
2. Males or females, ≥65 years of age.
3. Able to complete all scheduled visits and comply with all study procedures.
4. Women who were not of childbearing potential: surgically sterile (at least six weeks post bilateral tubal ligation, bilateral oophorectomy or hysterectomy); or postmenopausal (defined as a history of ≥12 consecutive months without menses prior to randomization in the absence of other pathologic or physiologic causes, following cessation of exogenous postmenopausal sex-hormonal treatment).
5. Participants in stable health in the opinion of the investigator for at least 30 days prior to screening (eg, no hospital admission for acute illness in the last 30 days prior to screening).

Exclusion Criteria

1. Participation or planned participation in an investigational clinical trial (eg, vaccine, drug, medical device, or medical procedure) within 30 days of Day 1 and for the duration of the study. **Note**: Participation in an observational trial or follow-up phase of a trial may have been allowed; however, these instances were to be discussed with the sponsor’s Medical Monitor (MM) prior to enrollment.
2. Prior receipt of any CHIKV vaccine.
3. Positive laboratory evidence of current infection with HIV (human immunodeficiency virus), HCV (hepatitis C virus), or HBV (hepatitis B virus).
4. Body mass index (BMI) ≥35 kg/m^2^.
5. History of any known or suspected allergy or history of anaphylaxis to any component of the IP.
6. History of any known congenital or acquired immunodeficiency or immunosuppressive condition that could impact response to vaccination (eg, leukemia, lymphoma, malignancy, functional or anatomic asplenia, alcoholic cirrhosis). **Note**: History of basal cell and squamous cell carcinoma of the skin or carcinoma in situ of the cervix considered cured was not exclusionary. History of a malignancy considered cured from over five years from the date of screening with minimal risk of recurrence was not exclusionary.
7. Prior or anticipated use of systemic immunomodulatory or immunosuppressive medications from six months prior to screening through Day 22. **Note**: For systemic corticosteroid use at a dose or equivalent dose of 20 mg of prednisone daily for 14 days or more within 90 days of screening through Day 22 was exclusionary. The use of inhaled, intranasal, topical, or ocular steroids was allowed.
8. Bleeding disorder or receipt of anticoagulants in the 21 days prior to screening, contraindicating IM vaccination, as judged by the investigator.
9. Moderate or severe acute illness with or without fever (oral temperature ≥100.4°F [≥38.0°C]).
10. Receipt or anticipated receipt of immunoglobulin from 180 days prior to screening through Day 22.
11. Medical condition (such as dementia) that, in the opinion of the investigator, could adversely impact the participant’s participation in or conduct of the study.
12. Evidence of substance abuse that, in the opinion of the investigator, could adversely impact the participant’s participation in or conduct of the study.
13. Identified as an investigator or employee of an investigator or study center with direct involvement in the proposed study, or identified as an immediate family member (ie, parent, spouse) of the investigator or employee with direct involvement in the proposed study.
14. Receipt or anticipated receipt of any vaccine from 30 days prior to Day 1 through Day 22.
15. Receipt or anticipated receipt of blood or blood-derived products from 90 days prior to screening through Day 22.
16. Any planned elective surgery that may have interfered with study participation or conduct.
17. Any other medical condition that, in the opinion of the investigator, could have adversely impacted the participant’s participation in or conduct of the study.

CHIKV human serum neutralizing antibody (SNA) assay

A Chikungunya virus neutralization assay (CHIKV human SNA assay) was used to measure serum neutralizing antibody titers to CHIKV in trial participants. The CHIKV human SNA assay is based on capacity of serum antibodies to neutralize recombinant CHIKV 181/25 virus (attenuated derivative of Asian strain AF15561) engineered to express luciferase (CHIKV-*luc*). Reductions of luciferase activity are quantified in cultures of Vero cells exposed to mixtures of CHIKV-*luc* virus and dilutions of test serum. Quantitation of reporter gene expression correlating to the level of CHIKV-*luc* virus infected cells, is determined by detection of luciferase activity in assay wells using luciferin substrate and a microplate luminometer to measure luminescence. The CHIKV antibody 80% neutralizing titer (NT_80_), calculated using linear regression and interpolation analysis, is the reciprocal of the maximum serum dilution that provides 80% protection of Vero cells from CHIKV-*luc* infection (80% reduction of luciferase activity compared to virus only control).

Statistical analysis

Coprimary endpoint: Day 22 seroresponse rate

The superiority of the immune response to CHIKV VLP vaccine over that to placebo was demonstrated at Day 22 by comparing seroresponse rates (the proportion of participants with a human SNA assay ≥100; considered the presumptive seroprotection rate) between the two treatment groups. The difference in seroresponse rates between the CHIKV VLP vaccine and placebo groups were calculated, along with the 95% CI for the difference based on the Newcombe hybrid score method. The lower bound of the two-sided 95% CI on the difference in seroresponse rates between CHIKV VLP vaccine and placebo groups had to be ≥70% (clinical significance). Additionally, the null hypothesis of no difference between seroresponse rate proportions was tested using a chi-square test with alpha=0.05 (statistical significance). No multiplicity adjustment was employed, and no covariate adjustment was performed.

The primary comparison was in the IEP across all age groups combined. Tests were repeated in the modified intent-to-treat (mITT) population as a measure of robustness, along with tests for each population in the separate age strata. No multiplicity adjustment was made for the analysis of the separate age strata as the primary population was the combined age groups.

Co-primary endpoint: Day 22 geometric mean titer

Day 22 GMTs were compared between CHIKV VLP vaccine and placebo treatment groups and were analyzed via a linear model based on an alpha=0.05. The primary model was an ANOVA, with logarithmically transformed anti-CHIKV SNA titers (log_10_) as the dependent variable and treatment group and trial site as the fixed effects in the model. The adjusted least square means and their 95% CIs calculated based on the ANOVA were back transformed and reported as the group GMT values. All tests were carried out at a two‑sided significance level of 0.05 and no adjustment for multiplicity was applied.

The primary comparison was in the IEP across both age groups combined. Tests were repeated in the mITT population as a measure of robustness, along with tests for each population in the separate age strata. No multiplicity adjustment was made for the analysis of the separate age strata as the primary population was the combined age groups.

Key secondary endpoint: seroresponse rate at Days 15 and 183, in that order

Seroresponse rates and seroresponse rate differences (CHIKV VLP vaccine minus placebo) with associated 95% CIs based on antibody titers measured at Days 15 and 183 were analyzed as described above for Day 22. For each time point, the null hypothesis of no difference between seroresponse rate proportions in the CHIKV VLP vaccine vs placebo group were tested using a chi‑square test with alpha=0.05.

Secondary endpoint: geometric mean titer at Days 15 and 183

For the comparison of CHIKV VLP vaccine to placebo, GMTs based on antibody titers measured at Days 15 and 183 were analyzed as described above for Day 22. Geometric mean fold increases for increase over Day 1 titer were analyzed as described for GMTs for each postvaccination timepoint.

Secondary endpoint: seroconversion at other titers

Secondary response rates at other titers (eg, ≥15 and 4-fold rise over baseline) were reported with associated two-sided 95% Wilson method CIs by scheduled visit for each treatment group using the IEP. The significance of the treatment group difference was assessed using chi-square tests at each visit for the IEP based on both age groups combined with an alpha=0.05.

Supplementary figures

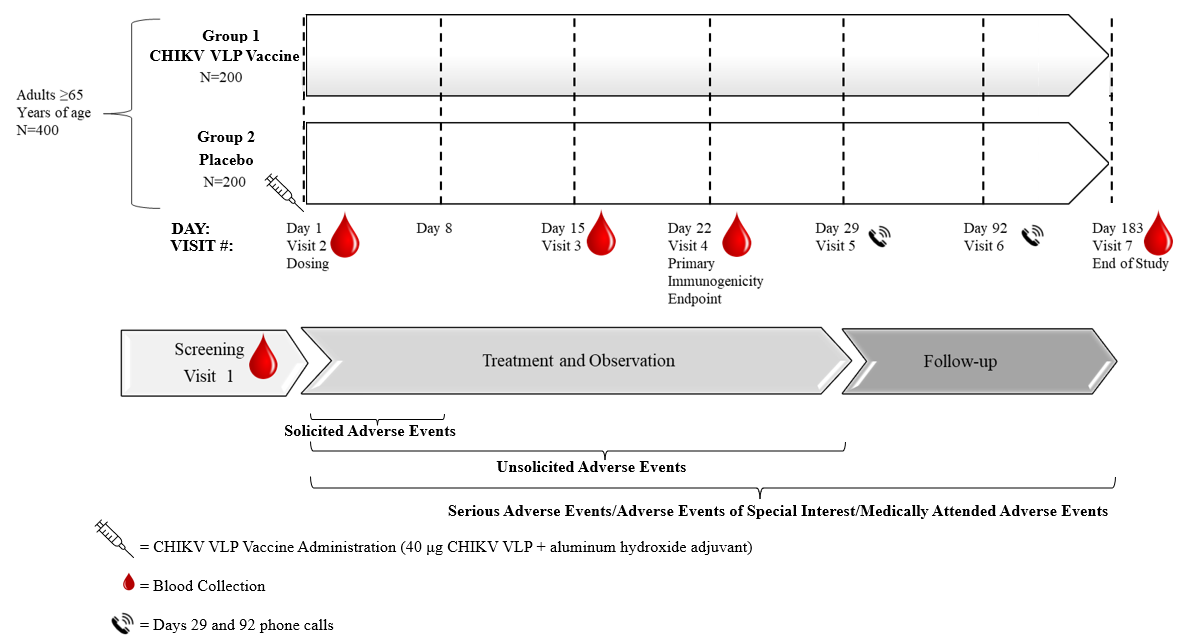

Figure S 1.	Trial Design.

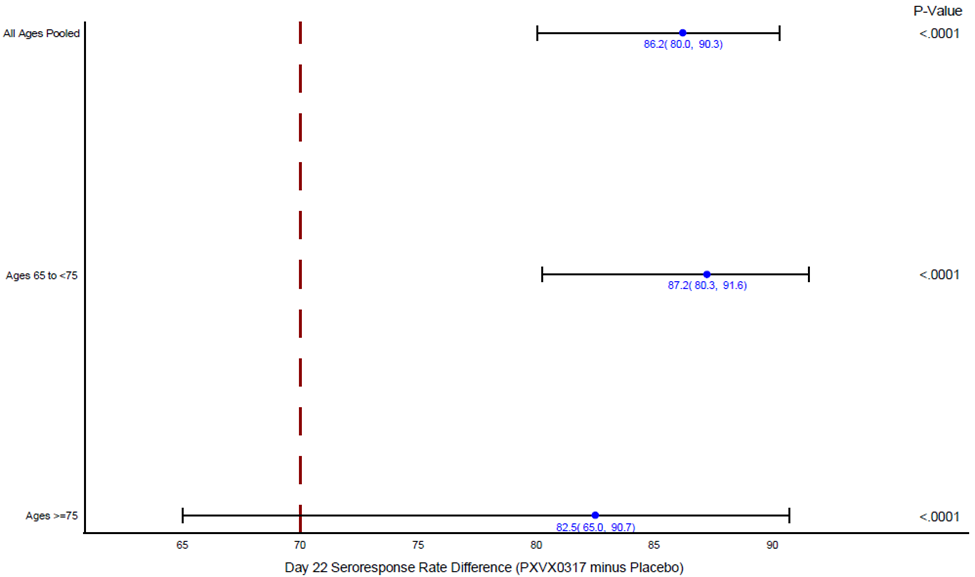

PXVX0317 = CHIKV VLP vaccine

Note: Error bars are 95% confidence interval based on the Newcombe hybrid score method; P values are from a two-sided chi-square test of equality of seroresponse percentages between groups.

Figure S 2. Forest Plot of Seroresponse Rate Differences at Day 22 (Immunogenicity Evaluable Population, All Ages Pooled and by Age Group).

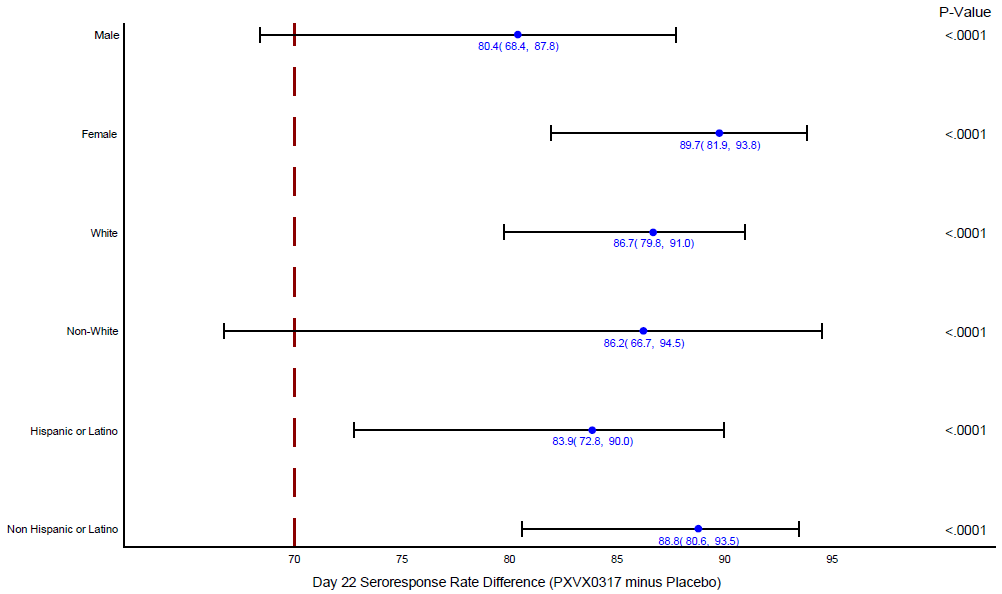

PXVX0317 = CHIKV VLP vaccine
Note: Error bars are 95% confidence intervals based on the Newcombe hybrid score method; P values are from a two-sided chi-square test of equality of seroresponse percentages between groups.

Figure S 3. Forest Plot of Seroresponse Rate Differences at Day 22 (Immunogenicity Evaluable Population, All Ages Pooled by Sex, Race, and Ethnicity Subgroups).

Supplementary tables

Table S 1	CHIKV SNA geometric mean titer and geometric mean fold increase from baseline by visit (immunogenicity evaluable population)

| Day  Statistic | CHIKV VLP Vaccine  (N=189) | Placebo  (N=183) | Ratio of GMTs  [95% CI]^a^ | P value^c,e^ |
| --- | --- | --- | --- | --- |
| **Day 1** | | | | |
| n^b^ | 189 | 183 |  |  |
| GMT^c^ [95% CI]  Median^d^  Min, Max^d^ | 7.50 [7.50, 7.50]  7.50  7.5, 7.5 | 7.50 [7.50, 7.50]  7.50  7.5, 7.5 | - | - |
| **Day 15** | | | | |
| n^b^ | 189 | 183 |  |  |
| GMT^c^ [95% CI]  Median^d^  Min, Max^d^ | 378.42 [301.04, 475.69]  499.80  7.5, 17062.5 | 8.96 [7.09, 11.32]  7.50  7.5, 9907.9 | 42.23 [32.13, 55.51] | <0.0001 |
| GMFI^e^ [95% CI]  Median^d^  Min, Max^d^ | 48.52 [40.12, 58.69]  66.64  1.0, 2275.0 | 1.16 [0.95, 1.40]  1.00  1.0, 1321.1 | - | <0.0001 |
| **Day 22** | | | | |
| n^b^ | 2559 | 424 |  |  |
| GMT^c^ [95% CI]  Median^d^  Min, Max^d^ | 723.93 [584.13, 897.20]  1009.60  7.5, 14518.9 | 8.08 [6.50, 10.04]  7.50  7.5, 1166.5 | 89.64 [68.62, 117.10] | <0.0001 |
| GMFI^e^ [95% CI]  Median^d^  Min, Max^d^ | 93.94 [78.03, 113.11]  134.61  1.0, 1935.9 | 1.05 [0.87, 1.27]  1.00  1.0, 155.5 | - | <0.0001 |
| **Day 183** | | | | |
| n^b^ | 2301 | 401 |  |  |
| GMT^c^ [95% CI]  Median^d^  Min, Max^d^ | 233.02 [194.05, 279.82]  284.25  7.5, 4091.2 | 8.29 [6.87, 10.02]  7.50  7.5, 193.9 | 28.09 [22.31, 35.38] | <0.0001 |
| GMFI^e^ [95% CI]  Median^d^  Min, Max^d^ | 29.32 [25.01, 34.37]  37.90  1.0, 545.5 | 1.04 [0.88, 1.22]  1.00  1.0, 25.9 | - | <0.0001 |
| CI = confidence interval; GMT = geometric mean titer; GMFI = geometric mean fold increase; LLOQ = lower limit of quantitation; Max = maximum; Min = minimum; SNA = serum neutralizing antibody.  Note: Values below lower limit of quantitation (LLOQ=15) are assigned the value LLOQ/2=7.5.  ^a^ Ratio of GMTs is (vaccine:placebo).  ^b^ n is the number of participants with a sample result available at the indicated visit.  ^c^ Geometric mean titer estimates, together with their 95% CIs, are derived from an ANOVA model that includes site and treatment group as fixed effects, assuming normality of the log titers. Ratio of GMTs and 95% CIs are derived from the same model. p-value tests equivalence of group GMTs on the log scale (ie, ratio of GMTs equal to 1).  ^d^ Median and range are based on raw values with imputation per note.  ^e^ Geometric mean fold increase from Day 1 (baseline) is (Day x/Day 1). GMFI estimates and 95% CIs are based on t-statistics assuming a normal distribution of the log fold increase in titer. P value tests equality of log fold increase in titer between groups. | | | | |

Table S 2	CHIKV SNA seroresponse rate by visit, sex, race, and age (immunogenicity evaluable population)

| Day | | Group | Subgroup | Seroresponse CHIKV VLP Vaccine (N=206)  **n/N (%)^c^ [95% CI]^d^** | Seroresponse Placebo (N=207)  **n/N (%)^c^ [95% CI]^d^** | Seroresponse Rate Difference  [95% CI]^a,b^ | P value |
| --- | --- | --- | --- | --- | --- | --- | --- |
| **Day 15** | | All IEP | All IEP | 149/181 (82.3) [76.1, 87.2] | 5/176 (2.8) [1.2, 6.5] | 79.5 [72.3, 84.6] | <0.0001 |
|  |  | Sex | Male | 51/70 (72.9) [61.5, 81.9] | 3/75 (4.0) [1.4, 11.1] | 68.9 [55.4, 78.3] | <0.0001 |
|  |  |  | Female | 98/111 (88.3) [81.0, 93.0] | 2/101 (2.0) [0.5, 6.9] | 86.3 [77.5, 91.3] | <0.0001 |
|  |  | Race | White | 123/151 (81.5) [74.5, 86.8] | 3/139 (2.2) [0.7, 6.2] | 79.3 [71.3, 84.9] | <0.0001 |
|  |  |  | Non-White | 26/29 (89.7) [73.6, 96.4] | 2/34 (5.9) [1.6, 19.1] | 83.8 [63.0, 91.8] | <0.0001 |
|  |  | Ethnicity | Hispanic or Latino | 63/77 (81.8) [71.8, 88.8] | 1/74 (1.4) [0.2, 7.3] | 80.5 [ 68.8, 87.6] | <0.0001 |
|  |  |  | Not Hispanic or Latino | 86/103 (83.5) [75.1, 89.4] | 4/104 (4.0) [1.6, 9.7] | 79.5 [69.4, 85.9] | <0.0001 |
|  |  | Age | ≥65 to <75 years | 121/144 (84.0) [77.2, 89.1] | 4/138 (2.9) [1.1, 7.2] | 81.1 [73.0, 86.5] | <0.0001 |
|  |  |  | ≥75 years | 28/37 (75.7) [59.9, 86.6] | 1/38 (2.6) [0.5, 13.5] | 73.0 [53.9, 84.2] | <0.0001 |
| **Day 22** | | All IEP | All IEP | 165/189 (87.3%) [81.8, 91.3] | 2/183 (1.1%) [0.3, 3.9] | 86.2 [80.0, 90.3] | <0.0001 |
|  |  | Sex | Male | 58/71 (81.7%) [71.2, 89.0] | 1/77 (1.3%) [0.2, 7.0] | 80.4 [68.4, 87.8] | <0.0001 |
|  |  |  | Female | 107/118 (90.7%) [84.1, 94.7] | 1/106 (0.9%) [0.2, 5.2] | 89.7 [81.9, 93.8] | <0.0001 |
|  |  | Race | White | 140/159 (88.1%) [82.1, 92.2] | 2/145 (1.4%) [0.4, 4.9] | 86.7 [79.8, 91.0] | <0.0001 |
|  |  |  | Non-White | 25/29 (86.2%) [69.4, 94.5] | 0/35 [0.0, 9.9] | 86.2 [66.7, 94.5] | <0.0001 |
|  |  | Ethnicity | Hispanic or Latino | 70/81 (86.4%) [77.3, 92.2] | 2/78 (2.6%) [0.7, 8.9] | 83.9 [72.8, 90.0] | <0.0001 |
|  |  |  | Not Hispanic or Latino | 95/107 (88.8%) [81.4, 93.5] | 0/104 [0.0, 3.6] | 88.8 [80.6, 93.5] | <0.0001 |
|  |  | Age | ≥65 to <75 years | 131/149 (87.9%) [81.7, 92.2] | 1/143 (0.7%) [0.1, 3.9] | 87.2 [80.3, 91.6] | <0.0001 |
|  |  |  | ≥75 years | 34/40 (85.0%) [70.9, 92.9] | 1/40 (2.5%) [0.4, 12.9] | 82.5 [65.0, 90.7] | <0.0001 |
| **Day 183** | | All IEP | All IEP | 139/184 (75.5) [68.9, 81.2] | 2/173 (1.2) [0.3, 4.1] | 74.4 [67.1, 80.1] | <0.0001 |
|  |  | Sex | Male | 43/70 (61.4) [49.7, 72.0] | 1/74 (1.4) [0.2, 7.3] | 60.1 [47.0, 70.7] | <0.0001 |
|  |  |  | Female | 96/114 (84.2) [76.4, 89.8] | 1/99 (1.0) [0.2, 5.5] | 83.2 [74.2, 88.8] | <0.0001 |
|  |  | Race | White | 119/155 (76.8) [69.5, 82.7] | 1/135 (0.7) [0.1, 4.1] | 76.0 [68.1, 82.0] | <0.0001 |
|  |  |  | Non-White | 20/28 (71.4) [52.9, 84.7] | 1/35 (2.9) [0.5, 14.5] | 68.6 [46.7, 82.1] | <0.0001 |
|  |  | Ethnicity | Hispanic or Latino | 60/77 (77.9) [67.5, 85.7] | 0/73 (0.0) [0.0, 5.0] | 77.9 [66.3, 85.7] | <0.0001 |
|  |  |  | Not Hispanic or Latino | 78/106 (73.6) [64.5, 81.0] | 2/99 (2.0) [0.6, 7.1] | 71.6 [61.1, 79.2] | <0.0001 |
|  |  | Age | ≥65 to <75 years | 112/147 (76.2) [68.7, 82.4] | 2/135 (1.5) [0.4, 5.2] | 74.7 [66.3, 81.0] | <0.0001 |
|  |  |  | ≥75 years | 27/37 (73.0) [57.0, 84.6] | 0/38 [0.0, 9.2] | 73.0 [54.6, 84.6] | <0.0001 |
|  | CI = confidence interval; IEP = immunogenicity evaluable population; SNA = serum neutralizing antibody  ^a^ Seroresponse rate difference is (vaccine minus placebo); 95% CIs are based on the Newcombe hybrid score method.  ^b^ P value is from a two-sided chi-square test of equality of seroresponse percentages between groups.  ^c^ n is the number of participants with seroresponse ≥ titer 100, divided by N, the total number of participants in the group.  ^d^ 95% CIs of seroresponse rates are based on the Wilson method. | | | | | | |

Table S 3	Participants with CHIKV SNA titer at or above selected titers (≥15 and 4-fold rise over baseline) by treatment group and visit (immunogenicity evaluable population)

| Day  Statistic | CHIKV VLP Vaccine  (N=189)  n (%) | Placebo  (N=183)  n (%) | Seroresponse Rate  Difference  ^[95%^ CI]^a^ | P value^b^ |
| --- | --- | --- | --- | --- |
| **Day 15** | | | | |
| n^c^ | 181 | 176 |  |  |
| n (%) titer ≥15  [95% CI]^d^ | 171 (94.5)  [90.1, 97.0] | 5 (2.8)  [1.2, 6.5] | 91.6 [86.0, 94.6] | <0.0001 |
| n (%) titer ≥ 4-fold Rise  [95% CI]^d^ | 155 (85.6)  [79.8, 90.0] | 5 (2.8)  [1.2, 6.5] | 82.8 [75.9, 87.5] | <0.0001 |
| **Day 22** | | | | |
| n^c^ | 189 | 183 |  |  |
| n (%) titer ≥15  [95% CI]^d^ | 180 (95.2)  [91.2, 97.5] | 2 (1.1)  [0.3, 3.9] | 94.1 [89.2, 96.5] | <0.0001 |
| n (%) titer ≥ 4-fold Rise  [95% CI]^d^ | 169 (89.4)  [84.2, 93.0] | 2 (1.1)  [0.3, 3.9] | 88.3 [82.4, 92.0] | <0.0001 |
| **Day 183** | | | | |
| n^c^ | 184 | 173 |  |  |
| n (%) titer ≥15  [95% CI]^d^ | 171 (92.9)  [88.3, 95.8] | 2 (1.2)  [0.3, 4.1] | 91.8 [86.3, 94.8] | <0.0001 |
| n (%) titer ≥ 4-fold Rise  [95% CI]^d^ | 153 (83.2)  [77.1, 87.9] | 2 (1.2)  [0.3, 4.1] | 82.0 [75.2, 86.8] | <0.0001 |
| Day  Statistic | | | | |
| **Day 15** |  |  |  |  |
| n^c^ | 181 | 176 |  |  |
| n (%) titer ≥15  [95% CI]^d^ | 171 (94.5)  [90.1, 97.0] | 5 (2.8)  [1.2, 6.5] | 91.6 [86.0, 94.6] | <0.0001 |
| CI = confidence interval; IEP = Immunogenicity Evaluable Population; SNA = serum neutralizing antibody.  Note: By definition, the IEP has no measurable CHIKV SNA at Day 1, therefore Day 1 is omitted from this table.  ^a^ Seroresponse rate difference is (vaccine minus placebo); 95% CIs are based on the Newcombe hybrid score method.  ^b^ p-value is from a two-sided chi-square test of equality of seroresponse percentages between groups.  ^c^ n is the number of participants with a sample result available at the indicated visit.  ^d^ 95% CIs of seroresponse rates are based on the Wilson method. | | | | |

Table S 4	Treatment-related unsolicited adverse events by preferred term and highest reported severity (safety population)

| **Preferred Term (MedDRA 24.1)** | CHIKV VLP Vaccine^a^ (N=206) | | | | Placebo^a^ (N=207) | | | |
| --- | --- | --- | --- | --- | --- | --- | --- | --- |
|  | Total  m, n (%) | Grade 1  m, n (%) | Grade 2  m, n (%) | Grade 3  m, n (%) | Total  m, n (%) | Grade 1  m, n (%) | Grade 2  m, n (%) | Grade 3  m, n (%) |
| Any Treatment-related Unsolicited AE | 6, 4 (1.9) | 3, 3 (1.5) | 2, 0 (0.0) | 1, 1 (0.5) | 7, 6 (2.9) | 6, 5 (2.4) | 1, 1 (0.5) | 0 |
| Fatigue | 1, 1 (0.5) | 0 | 0 | 1, 1 (0.5) | 2, 2 (1.0) | 2, 2 (1.0) | 0 | 0 |
| Myalgia | 1, 1 (0.5) | 0 | 1, 1 (0.5) | 0 | 1, 1 (0.5) | 1, 1 (0.5) | 0 | 0 |
| Arthralgia | 1, 1 (0.5) | 0 | 1, 1 (0.5) | 0 | 0 | 0 | 0 | 0 |
| Diarrhoea | 1, 1 (0.5) | 1, 1 (0.5) | 0 | 0 | 0 | 0 | 0 | 0 |
| Feeling abnormal | 1, 1 (0.5) | 1, 1 (0.5) | 0 | 0 | 0 | 0 | 0 | 0 |
| Hypertension | 1, 1 (0.5) | 1, 1 (0.5) | 0 | 0 | 0 | 0 | 0 | 0 |
| Pruritus | 0 | 0 | 0 | 0 | 1, 1 (0.5) | 0 | 1, 1 (0.5) | 0 |
| Chills | 0 | 0 | 0 | 0 | 1, 1 (0.5) | 1, 1 (0.5) | 0 | 0 |
| Musculoskeletal discomfort | 0 | 0 | 0 | 0 | 1, 1 (0.5) | 1, 1 (0.5) | 0 | 0 |
| Rhinorrhoea | 0 | 0 | 0 | 0 | 1, 1 (0.5) | 1, 1 (0.5) | 0 | 0 |
| *There were no grade 4 treatment-related unsolicited AEs.  AE = adverse event; Grade 1 = mild; Grade 2 = moderate; Grade 3 = severe; m=Number of events, n=Number of participants with events  Note: Percentages are based on the number of safety population participants in each treatment group. Participants are counted once within each preferred term. Preferred terms are ordered by descending frequency within the vaccine group. | | | | | | | | |
